# Advanced epithelial ovarian cancer in the older patient: a retrospective cohort study of six UK Gynaecological Cancer centers

**DOI:** 10.1101/2025.04.03.25325200

**Authors:** Victoria Cullimore, Kezia Gaitskell, Rebecca Newhouse, Kathryn Baxter, Nicholas Wood, Christina Fotopoulou, Jason Yap, Madeline MacDonald, Richard J Edmondson, Jo Morrison

## Abstract

**Objective:** We aimed to analyze management and survival outcomes of older people (≥75 years) with stage II+ epithelial ovarian cancer (EOC) across six gynaecological cancer centers in the United Kingdom.

**Methods:** Retrospective cohort study performed using the IMPRESS project dataset. Clinical information for patients diagnosed with EOC from six sites of varying size and population demographics, was collated between 1/1/2018 and 31/12/2019. We compared treatment of patients aged ≥75 years with those <75, within and between centers, using multi-variate analysis, to understand effects on outcomes.

**Results:** After exclusions, we assessed 721 people for overall survival (OS) and 702 for progression-free survival (PFS). The ≥75s had poorer performance status, more comorbidities and were less likely to receive combination treatment with surgery and chemotherapy (in either order) (overall = 392/721 (54.4%) overall; <75 cohort = 320/495 (64.6%); ≥75 cohort = 72/226 (31.9%). Treatment proportions varied between sites, with some having no active treatment rates of nearly 50% for ≥75s. Those ≥75 years had twice the relative risk of death compared to those <75 (Relative Risk (RR) = 1.98, 95% confidence intervals (CI) 1.63 to 2.39, P<0.001). Adjustment for confounders individually caused only a relatively modest reduction in magnitude and strength of association. Adjustment for treatment led to this association essentially disappearing (RR = 1.10, 95% CI 0.88 to 1.38; 99% reduction in Chi2), though with significant variation in association between age and OS between treatment groups (p-heterogeneity: 0.0004).

**Conclusion:** Older women may do as well as younger women in terms of survival, if treated similarly, although this varies depending on treatment groups. Treatments varied between and within sites, with some sites treating older women more differently than others. Some differences may be appropriate, but significant differences in rates of no active treatment between sites suggests that not all variation may be appropriate.

**Key messages:** What is already known on this topic?

- Older patients with ovarian cancer are under-represented in clinical trials.
- Optimal management of advanced ovarian cancer requires a combination of surgery and chemotherapy.
- Management of older patients remains a challenging area due to the lack of consensus regarding optimal treatment strategies tailored to this population.
- Evidence suggests that older patients often receive less aggressive treatment than younger counterparts, partly due to concerns about comorbidities and treatment tolerance, despite similar attitudes to treatment.

**What this study adds?:**

- Treatment of older patients varied between and within centers.
- A greater percentage of older patients died within 30 or 90 days of diagnosis.
- Older patients were less likely to have combination treatment with surgery and chemotherapy, especially in some centers.
- Older patients who had combination treatment had similar outcomes to the younger cohort.

**How this study might affect research, practice or policy?:**

- Further work is needed to understand which older patients will do better if treated with surgery and/or chemotherapy and in which order, and to improve shared decision-making with our patients.
- This study suggests that age alone should not be a barrier to enrolment in clinical studies and active recruitment of older people should be encouraged.

## Introduction

Epithelial ovarian cancer (EOC) is the leading cause of gynaecological cancer-related mortality.^1^ Globally in 2022, 324,398 females were diagnosed with ovarian cancer.^2 3^ EOC predominantly affects older females, with peak incidence in 75 to 79-year-olds^3^ with incidence anticipated to rise as populations age.^4 5^ Survival rates significantly differ by age; in the United Kingdon (UK) only 21.5% of patients diagnosed with ovarian cancer over 75 years survive for over ten years, in contrast to 81.2% of those aged 15 to 44 years.^3^ The global population is ageing; by 2050 the number aged 80 years or older is expected to triple, from 157 million in 2022 to 459□million by 2050,^6^ with ovarian cancer already the third most common cause of cancer-related mortality in females over 80 years of age in the Indian subcontinent.^7^

Current guidelines for EOC recommend a combination of chemotherapy and cytoreductive surgery for advanced disease (FIGO stages III and IV).^8^ Cytoreduction to leave no macroscopic residual disease (NMRD), is associated with both increased overall (OS) and progression-free survival (PFS).^9^ Standard of care may consist of primary cytoreductive surgery (PCRS), followed by platinum-based chemotherapy, or neoadjuvant chemotherapy (NACT) with interval cytoreductive surgery (ICRS), which has been shown to be non-inferior. ^10 11^ NACT/ICRS should be considered for patients in whom maximal primary surgical effort may not be tolerated or NMRD achieved at PCRS.^5^

Several factors may contribute to the differential management of older patients with ovarian cancer, including aggressive histological subtypes, sub-optimal treatment, lower physiological reserve, delayed or emergency presentations and increased toxicity.^12 13^ Additionally, co-morbidities are more prevalent in older people, correlating with increased mortality and surgical complications in older patients with EOC.^14^ However, treatment more frequently deviates from the optimal approach in older people and the probability of receiving standard treatment in those aged over 70 years is around 50%.^15^ People over 65 years are more likely to undergo less-radical surgery, resulting in higher residual disease, and typically receive fewer cycles of both adjuvant and neoadjuvant chemotherapy.^15-17^ In England, between 2016 and 2018, 60% of patients over the age of 79 had no record of *any* treatment for their ovarian cancer.^18^ A study of 1123 people with EOC found that the proportion of people with residual disease was 30% in older cohorts, compared to 20% in the under 65s, and only 40% of people aged 75 and over underwent a standard procedure, compared to 72% of those aged under 65.^17^ Alongside less radical surgery, fewer older patients receive chemotherapy, often given a lower dose or a non-standard regimen, such as platinum monotherapy.^12^

The optimal treatment strategy in the older population remains unclear, as they are underrepresented in clinical trials of cancer management.^19^ Personalization of cancer care is essential for all patients and a comprehensive geriatric assessment prior to treatment, incorporating functional dependence, comorbidity, organ function and cognitive and psychosocial factors, may enhance management decisions.^4^

This was a sub-study of data collected for the IMPRESS project, ^20^ analyzing management and outcomes of people over the age of 75 years, treated for stage II-IV epithelial ovarian cancer in six gynaecological cancer centers in the United Kingdon (UK). We wanted to understand how outcomes varied for older people, with adjustment for other risk factors, and how treatments differed within and between centers.

## Materials and methods

The IMPRESS project collected data from six sites in the UK of varying size, geography and surgical resection rates.^20^ Data were collected retrospectively for all patients treated for ovarian cancer between 1/1/2018 and 31/12/2019.

A data dictionary was used to document demographic data encompassing ethnicity, index of deprivation and background clinical information including BMI, co-morbidities and performance status. A secure web-based database (REDCap) was used for data collection, which included method of diagnosis, histological sub-type, grade and stage, surgical and oncological management, recurrence and survival.

We chose to split at age 75 years, since there were relatively few people aged over 80. Patients were excluded if data on date of diagnosis, survival status, or recurrence status were missing. Patients with early-stage disease (stage I) and with non-epithelial or low-grade serous carcinomas were excluded.

For full methodological details see Supplementary Materials. Briefly, descriptive tabulations with Chi-squared (Chi-2) tests for association were used to explore variation in patient and tumour characteristics and patterns of treatment between people diagnosed with EOC aged <75 versus ≥75. Overall survival (OS) and progression-free survival (PFS) were examined using Kaplan-Meier analysis for survival curves and Cox models for regression analysis, with time since diagnosis as the underlying time variable. Cox regression models were used to estimate hazard ratios (hereafter referred to as relative risks (RR)) for the association between age at diagnosis (≥75 versus <75) and survival with adjustment for potential confounders. Stratification was used as an alternative to adjustment where there was concern regarding violation of the non-proportional hazards assumption.

Ethical approval was granted from the HRA and Health and Care Research Wales for data collection and storage (ref 22/HRA/3264). In accordance with the journal’s guidelines, we will provide our data for independent analysis by a selected team by the Editorial Team for the purposes of additional data analysis or for the reproducibility of this study in other centers, if such is requested.

## Results

Data were collected for 1117 women in total. For this study, all patients with non-epithelial ovarian cancer (n= 25), low grade serous cancer (n = 47) and low-stage disease (n=247) at diagnosis were removed before analysis. Seventy-seven patients were excluded due to missing survival status or other essential information, retaining 721 patients for analysis of overall survival and 702 for recurrence-free survival (19 patients excluded due to unknown recurrence information) (Figure 1).

### Baseline characteristics

Overall, 226/721 (31%) of the cohort were aged ≥75 years (Table 1), although this differed significantly between centers with some having as low as 18% (Site 6) aged ≥75 and others up to 39% (Site 1) (p < 0.001) (Supplementary Table S1). Ethnicity varied significantly between <75 and ≥75 cohorts, with the older cohort being less diverse (p = 0.008). The older cohort was also less likely to smoke (p < 0.001) and was thinner (p < 0.001), poorer performance status (p < 0.001), and higher levels of co-morbidity (p = 0.005), although there was no difference between groups in terms of deprivation index (p = 0.3).

**Table 1.**
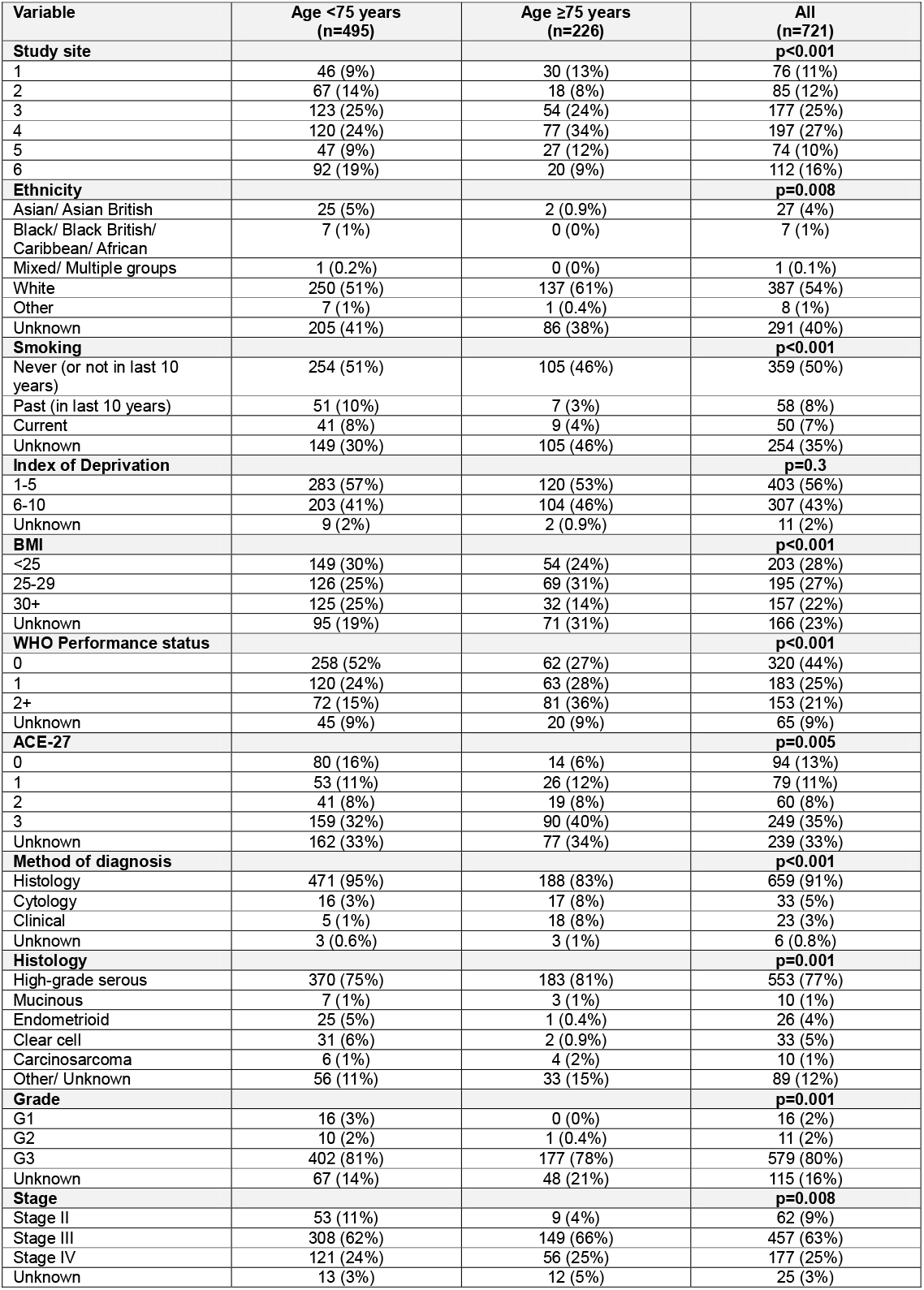
Demographic and diagnostic variables by age at diagnosis <75 vs. ≥75 years. Table demonstrates column %; P-value is for Chi-2 test for association between variable and age.

Younger patients were more likely to have non-high grade serous pathology, and older patients were more likely to have histological subtype and grade not recorded (p = 0.001) (Table 1). Diagnosis based either on clinical factors or cytology was also more likely in the older cohort (p <0.001), perhaps indicating poorer performance status, less primary surgery and/or unwillingness to put older patients through invasive procedures. Patients ≥75 were more likely to present with more advanced disease (stage III+ or unknown: <75 = 442/495 (89%); ≥75 = 217/226 (96%), p = 0.008).

### Treatment

Older patients were less likely to be treated with a combination of chemotherapy and surgery (See Figure 2, Supplementary Figure S1, and Supplementary Table S2). Unfortunately, mode of treatment was unclear for 88/721 people (12%), mostly from Site 3, due to reliance on cancer registry data without additional data cleaning. If these women were excluded, 320/441 (73%) women in the <75 cohort had a combination of surgery and chemotherapy, compared with 72/192 (38%) women in the ≥75 cohort, and 56/192 (29%) in the ≥75 cohort had no active cancer treatment, compared with just 22/441 (5%) in the <75 cohort. Overall, even including the ‘other’ treatments, those ≥75 years with Stage II+ EOC appear to be treated differently to younger women (Figure 2 and Supplementary Figure S1). In particular, a smaller proportion of older women received combined chemotherapy and surgery (overall = 392/721 (54%) overall; <75 cohort = 320/495 (65%); ≥75 cohort = 72/226 (32%)), and a higher proportion received neither chemotherapy nor surgery (overall = 88/721 (12%) overall; <75 cohort = 22/495 (4%); ≥75 cohort = 56/226 (25%)) (See Table S2).

Among those who received combination therapy (surgery and chemotherapy in either order), characteristics were similar between those aged <75 and those aged ≥75. Unsurprisingly, the subset of older women who received combination treatment had different characteristics to the total cohort of older women (e.g., PS = 0 in 62/226 (27%) of those aged ≥75 overall, but in 33/72 (46%) of those aged ≥75 who received combined treatment) (Table 1 and Supplementary Table S3). Numbers of patients who received no treatment were small, especially for those aged <75, which limits conclusions. However, those who did not receive any treatment were typically of poor performance status ^21^, had more comorbidities^22^, and high stage disease, regardless of age.

Overall, taking all ages together, treatment varies between centers (see Figure 2 and Supplementary Table S4). In some centers, there is also variation in treatment by age (see Figure 2 and Supplementary Table S5). Some of this may be due to differences in populations and referral patterns, since some of the sites take more tertiary referrals and see fewer patients who are referred to them directly, compared to others.

This may partly explain why two sites entered 0-1% of patients who had no active treatment, although missing data for older patients is a more likely explanation, from recent National Ovarian Cancer Audit data.^23^

### Survival

#### Overall survival

The mean follow-up time for overall survival analyses was 1.90 years (standard deviation: 1.37). Without adjustment, women with EOC aged ≥75 had approximately twice the relative risk of death compared to patients aged <75 (RR = 1.98, 95% confidence interval (CI) 1.63 to 2.39, P<0.001; Figure 3 and Supplementary Figure S2). Deaths in those aged ≥75 were particularly noticeable soon after diagnosis, with 7.5% dying within 30 days (compared to 1.0% of those aged <75), and 22% dying within 90 days (compared to 7.1% of those aged <75) (see Figure 3). Adjustment for most potential confounders individually caused only a relatively modest reduction in the magnitude and strength of the association (as assessed by the size of the hazard ratio and the percentage change in the Chi-2; Supplementary Figure S2). However, adjustment for treatment led to this association essentially disappearing (RR = 1.10, 95% CI 0.88 to 1.38; 99% reduction in Chi-2). When potential confounders were dealt with by stratification (to allow for potentially non-proportional hazards), results were similar (Supplementary Figure S3). Multivariable Cox regression models for the association between age and survival included study site, stage, and histological type as a-priori confounders; performance status, smoking, BMI, method of diagnosis, and treatment were also included as adjustment/stratification factors as they caused a >10% change in the Chi-2 statistics for association between age and survival (see Supplementary Figures S2 and S3).

On visual inspection of the Kaplan-Meier survival curves, the association between age and overall survival appeared to vary substantially between treatment groups (Figure 4). This is supported by adjusted Cox regression models showing strong evidence of statistical heterogeneity in the association between age and OS by treatment group (likelihood ratio test for a multiplicative interaction term between age and treatment, p-heterogeneity = 0.0004) (Supplementary Figure S4). Women aged ≥75 at diagnosis have substantially worse survival than those aged <75 in the groups treated with surgery alone (RR = 16.8, 95% CI 4.11 to 68.27) or with other/unclear treatment regimens (RR = 2.83, 95% CI 1.35 to 5.96), though small numbers in some groups led to wide confidence intervals. The difference in survival is more modest or absent in those treated with chemotherapy alone (RR = 1.16, 95% CI 0.72 to 1.88) or with some combination of chemotherapy and surgery (chemotherapy then surgery, RR = 1.79, 95% CI 1.07 to 2.99); surgery then chemotherapy, RR = 1.17, 95% CI 0.53 to 2.59). By contrast, survival appears worse in the younger group in those receiving neither surgery nor chemotherapy, though this is not statistically significant (RR = 0.54, 95% CI 0.25 to 1.17) (Figure 4, Supplementary Figure S4).

The Kaplan-Meier survival graphs for OS by study site, performance status, stage at diagnosis, and histological type appear marginally better for women aged <75 versus ≥75, though small numbers in some groups make comparisons challenging (Supplementary Figure S5).

### Progression-free survival

In unadjusted analyses, there was no evidence of an association between age at diagnosis and PFS (comparing age ≥75 versus <75, RR = 0.88, 95% CI 0.66 to 1.18, p=0.4). However, as with OS, there was evidence of variation in the association between age and PFS between treatment groups (p-heterogeneity = 0.03), with a similar pattern of associations to OS (Supplementary Figures S6 and S7).

## Discussion

### Summary of Main Results

These data demonstrate that older patients do not necessarily have worse disease and do as well as younger women in terms of survival, if able to be treated similarly with certain treatment regimens (for example, surgery then chemotherapy). However, treatments vary between centers, with some centers treating older people more differently than others. Some of the differences in treatment are appropriate, due to comorbidities and frailty, but the significant differences in rates of no active treatment between sites, and associated differing survival rates, suggests that not all this variation may be appropriate.

### Results in the Context of Published Literature

The IMPRESS data demonstrate that in many centers, older patients with EOC receive different treatment compared with younger patients. This is in line with previous studies, although these have not compared the effect of different centers, with differing attitudes to ovarian cancer treatment, to outcomes. Rousseau at al attributed differences to more aggressive histological subtypes, less willingness to consider surgery and/or chemotherapy and delays and reduced doses of chemotherapy.^12^ Others have found that older people presented later with higher stage disease, had lower rates of achieving NMRD at cytoreduction, if offered, were more likely to have surgery with non-specialists in smaller centers and less likely to be offered standard chemotherapy regimens.^17^ Another study reported that only half of patients aged over 65 with advanced EOC were treated with platinum-based chemotherapy. However, they found that survival improved by 38% in those treated with platinum-based chemotherapy, similar in magnitude to the benefits described in RCTs (which recruit younger cohorts).^24^ There was greater improvement to those given platinum agents in combination with paclitaxel. Generally, the literature supports the theory that older patients have poorer outcomes and much of this is due to reduced access and radicality of treatment.

However, a retrospective study by Mallen et al concluded that comorbidities had less effect than tumour biology, due to lower rates of BRCA mutations and lower rates of platinum sensitivity.^25^ However, they noted that older patients (>70 years) also had lower rates of: enrollment in clinical trials; completion of adjuvant chemotherapy; intraperitoneal chemotherapy; surgery resulting in NMRD. This is despite the willingness of older patients to have radical treatment and valuing cure as much as younger patients, and older patients not perceiving age as a barrier to treatment.^26 27^

### Strengths and Weaknesses

A weakness of this study was that we collected data from only six centers. However, these were chosen from the National Ovarian Cancer Audit Feasibility Pilot to cover a spectrum of locations, sizes and surgical practices, so are likely to be a representative sample of the national picture.^18^ Sites ranged from most likely out of 40 gynaecological oncology cancer centers in England to receive any treatment, to 36/40 in the recent National Ovarian Cancer Audit (NOCA) for 2022 and its associated data.^23^ The depth of data collected for the IMPRESS study was extensive, with much greater granularity than normally available from population-based studies. Data quality from five of the sites was good, although one site uploaded data from their cancer registry without extensive data cleaning, so information about treatment types etc., was unclear for a proportion of their patients.

Sites were expected to collect data for all patients with ovarian cancer in the time-period, although two centers had much lower numbers than one would have expected from their population sizes and annual data from the NOCA.^23^ This raises concerns about selective reporting bias for these sites, especially as no treatment rates were 0-1% in two centers in this dataset, compared with 31.1% and 11.2%, respectively, in the most recent NOCA dataset.

One of the challenges in interpreting these data is that treatment received will be inevitably (and appropriately) affected by patient fitness/frailty and extent of disease. While we have attempted to adjust for these, the measures available and consequent adjustment will be imperfect. Thus, for example, the observed very poor survival in women aged <75 who received neither chemotherapy nor surgery is likely to reflect the fact that, in younger women, only those with very poor fitness or very end-stage disease would not receive treatment.

### Implications for practice and future research

These data suggest that older people with similar degrees of fitness, comorbidities and disease have broadly similar survival rates, if treated similarly to younger people with EOC. In those treated with combination of surgery and chemotherapy, in either order, survival rates for the first 18 months were identical, then separated. This perhaps reflects effects of age and comorbidities but may also reflect a lower likelihood of second line treatment at recurrence, since PFS rates were identical. Other studies have shown increased mortality in ≥75s within 30-days of surgery,^28^ so careful shared decision-making is required to assess the risks and benefits of either approach. Patient decision-aids and improved shared decision-making may support patients to make choices for themselves as individuals. This requires people to be informed of available options and the risks and benefits of the different options, as priorities differ between patients, and between patients and clinicians.^29^ It is in these important conversations that we meld the art and science of medicine.^30^ Improved shared decision-making should help to reduce ‘group think’ in MDTs,^31^ and, we hope, improve outcomes by reducing variability.

Data from studies show that comorbidity is associated with mortality and surgical morbidity in older women with ovarian cancer.^14^ A position paper highlighted the need for multi-disciplinary, including onco-geriatric, assessment in older people with ovarian cancer.^32^ Increased use of co-management with oncologists, surgeons and care of the older person specialists may improve outcomes for older people with ovarian cancer, but is yet to be proven by RCTs,^7 33 34^ although a study is ongoing.^35^ Current data, including these in this study suggest that age alone should not be a barrier to effective treatment, but inclusive decision-making is required to avoid unnecessary harm.

## Data Availability

we will provide our data for independent analysis by a selected team for the purposes of additional data analysis or for the reproducibility of this study in other centers, if such is requested, if in accordance with the ethical approval requirements.

## Acknowledgements

We are grateful to all at the sites involved in data collection for the IMPRESS study^20^ and thank them for their candor and willingness to be involved in a project that aims to reduce variability and improve care for women with ovarian cancer. We thank Ovarian Cancer Action who funded the IMPRESS study and those at the participating centers for their contribution to data collection.

## Authors’ contributions

JM conceptualized this study. RJE, KB, JM, NW, CF, JY and MMcD designed and conducted the IMPRESS study, which provided the dataset used for the analysis in this study. VC and RN contributed to the original IMPRESS data collection. VC, RN, KG and JM designed this study. Statistical analysis was performed by KG and data analyzed by KG, VC, RG and RJE. VC, KG and JM wrote the first draft of the manuscript. All authors contributed to revisions and have approved the final version. JM acts as guarantor.

---

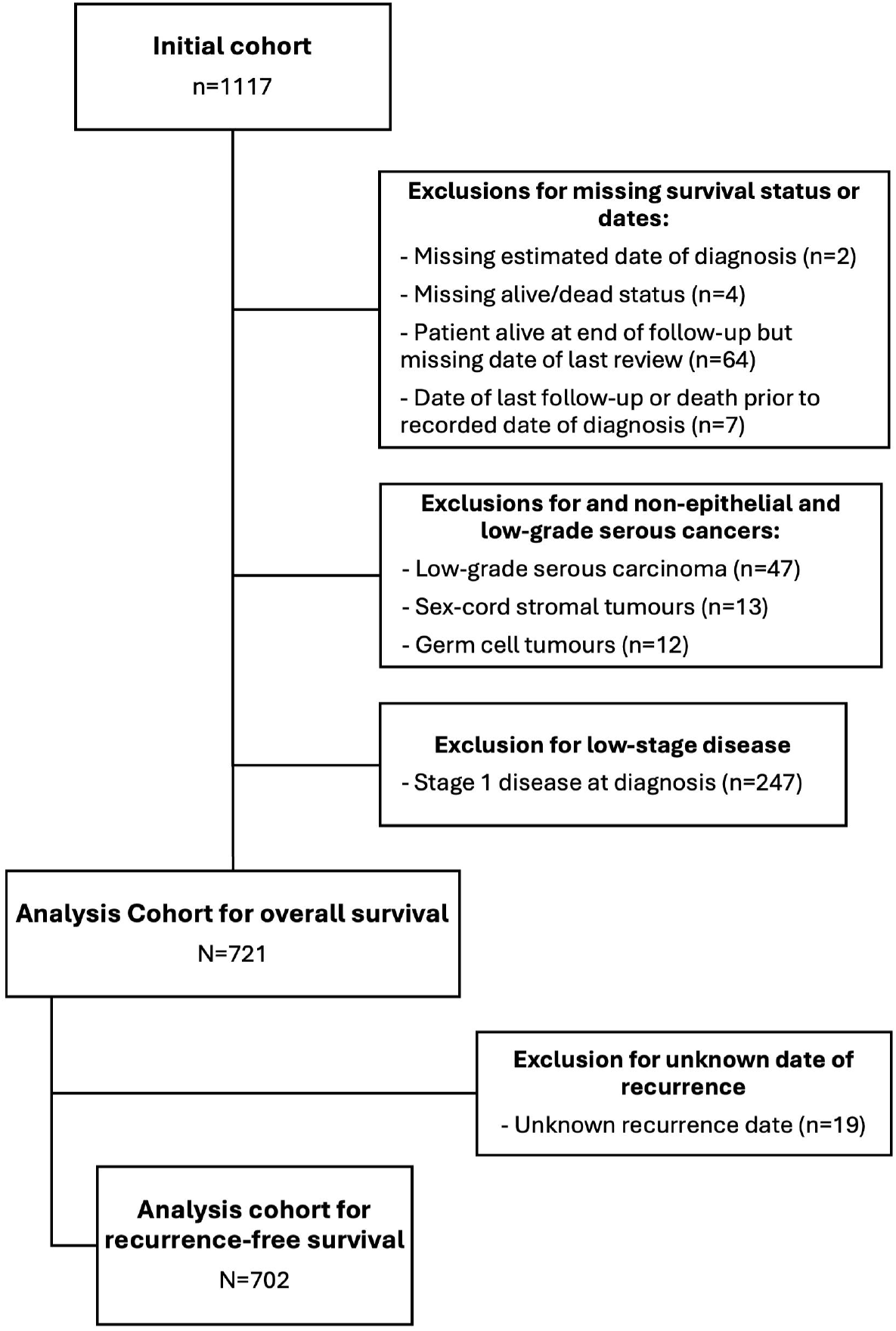

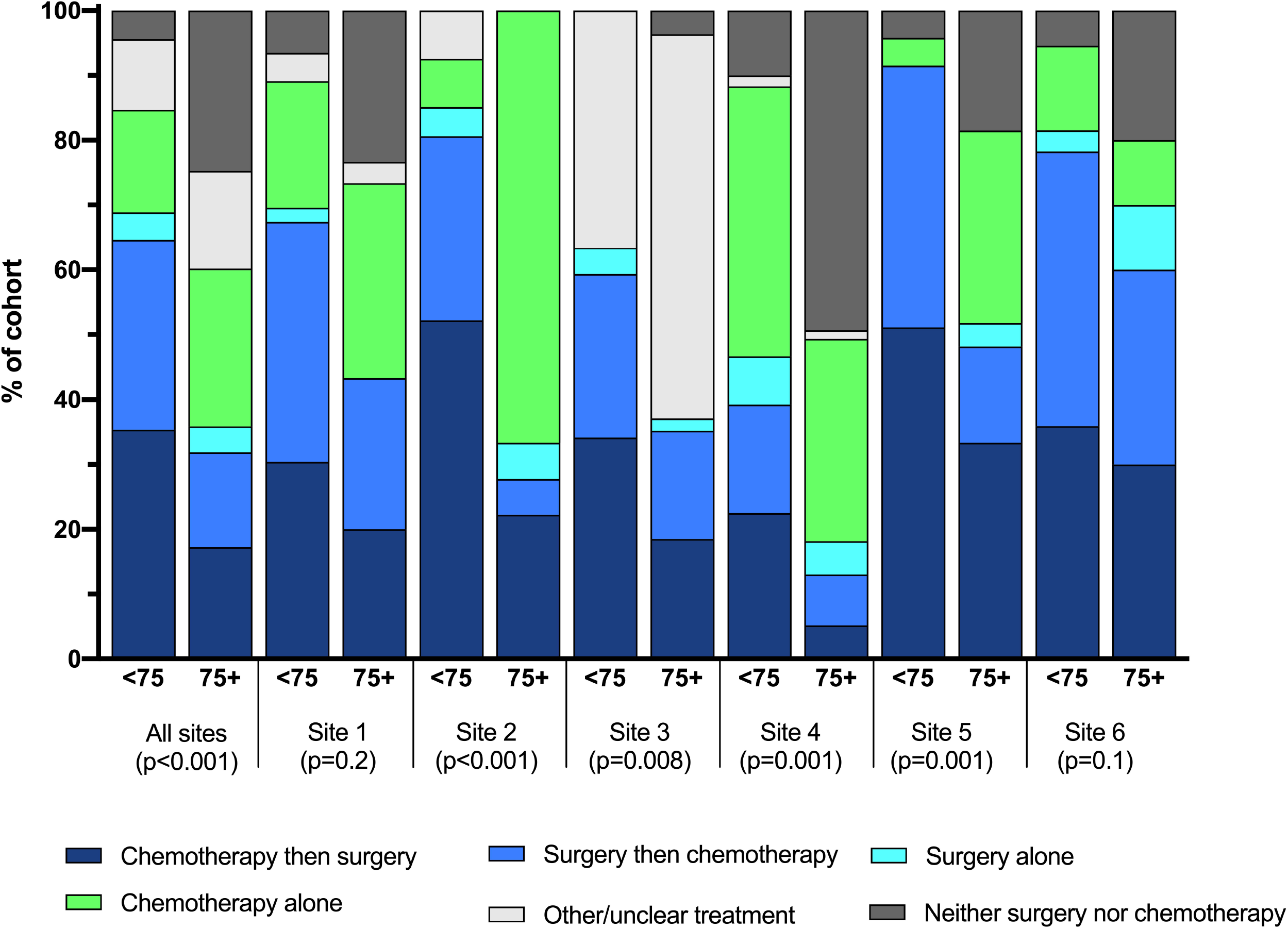

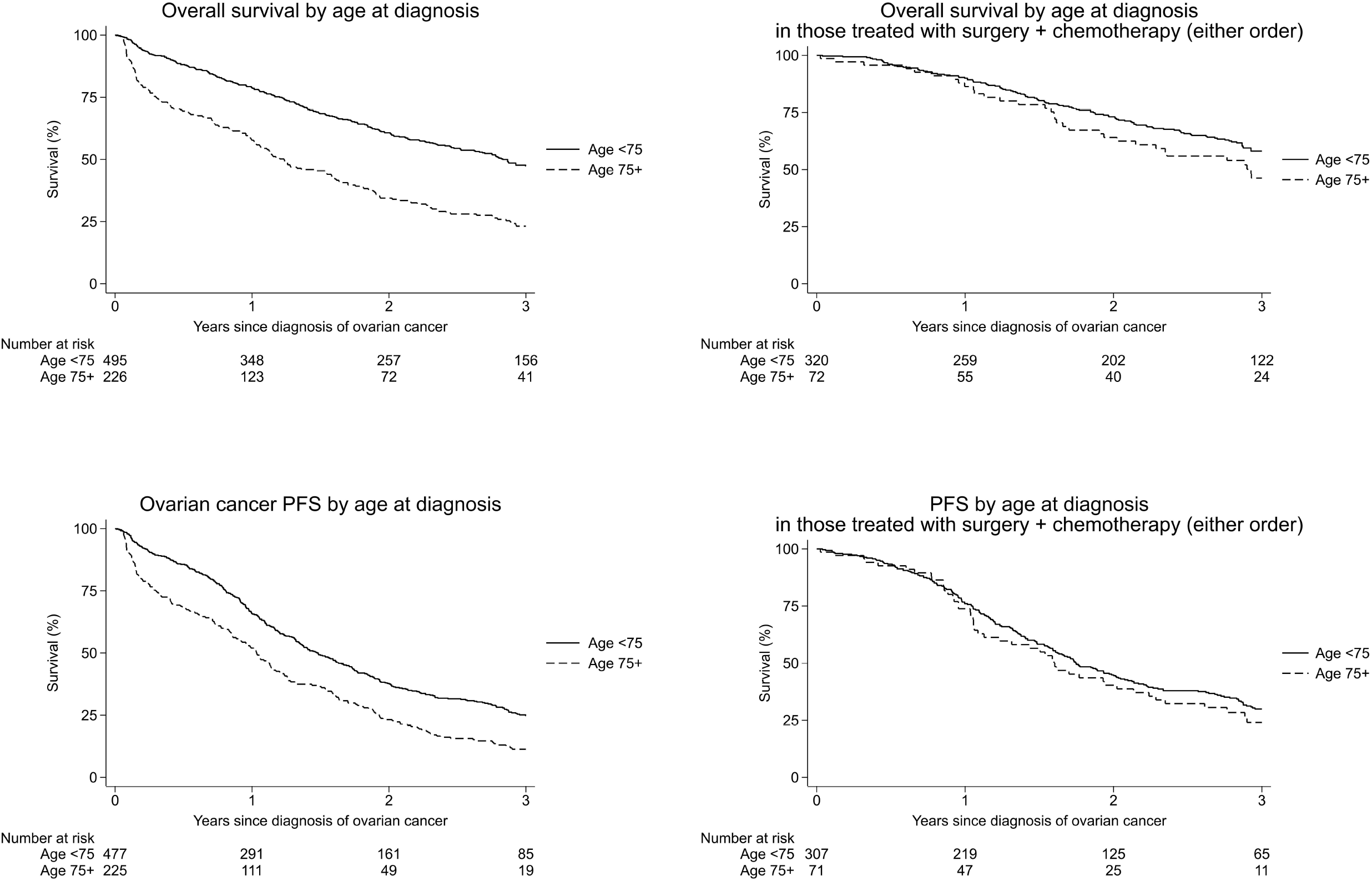

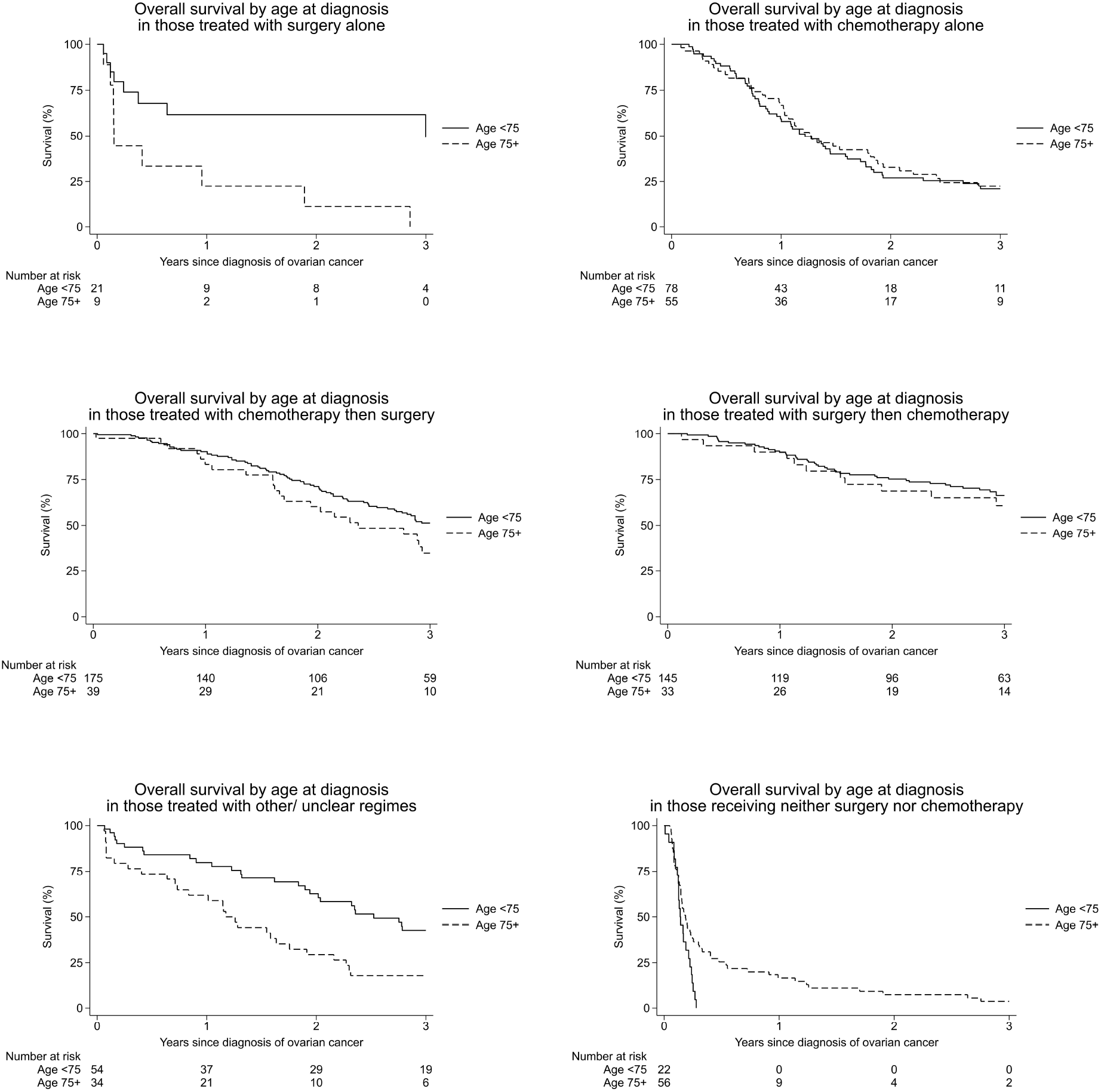

