## Supplementary Materials for "Advanced epithelial ovarian cancer in the older patient: a retrospective cohort study of six UK Gynaecological Cancer centers"

| **Supplementary information** |  |  |
| --- | --- | --- |
| Methods details | Additional detail on methods |  |
| **Supplementary Tables** |  |  |
| Table S1 | Age distribution of patients with ovarian cancer by study site | 3 |
| Table S2 | Treatment of patients with ovarian cancer by age at diagnosis | 3 |
| Table S3 | Patient characteristics by age at diagnosis in those who received combination therapy (chemotherapy + surgery, in either order) | 4 |
| Table S4 | Treatment for ovarian cancer by study site | 5 |
| Table S5 | Treatment for ovarian cancer by study site and age at diagnosis | 5 |
| **Supplementary Figures** |  |  |
| Figure S1 | Sankey diagram showing differences in treatment pathways for ovarian cancer by age at diagnosis | 6 |
| *Overall survival* |  |  |
| Figure S2 | Association between age at diagnosis (75+ versus <75 years) and overall survival (risk of death) with adjustment for various factors | 7 |
| Figure S3 | Association between age at diagnosis (75+ versus <75 years) and overall survival (risk of death) with stratification for various factors | 8 |
| Figure S4 | Association between age at diagnosis (75+ versus <75 years) and overall survival (risk of death) by treatment group | 9 |
| Figure S5 | Kaplan-Meier graphs of overall survival (OS) by study site, performance status, stage at diagnosis, and histological type, shown separately for women aged <75 years versus 75+ years at diagnosis | 10 |
| *Progression-free survival* | | |
| Figure S6 | Kaplan-Meier graphs of progression-free survival (PFS) by age at diagnosis and treatment group | 11 |
| Figure S7 | Association between age at diagnosis (75+ versus <75 years) and progression-free survival (risk of progression or death) by treatment group | 12 |

**Additional detail on methods**

Descriptive tabulations with Chi-squared (Chi-2) tests for association were used to explore variation in patient and tumour characteristics and patterns of treatment between women diagnosed with ovarian cancer aged <75 versus ≥75. Patient characteristics included: study site (six sites); ethnicity (Asian/ Asian British, Black/ Black British/ Caribbean/ African, Mixed/ Multiple groups, White, Other); smoking (never (or not in last 10 years), past (in last 10 years), current); Index of Multiple Deprivation (IMD) (10 categories based on area-level deprivation ^1^); body mass index (BMI) (<25, 25-29, 30+ kg/m^2^); WHO performance status ^2^ (0, 1, 2+); ACE-27 ^3^) (Adult Comorbidity Evaluation-27 (0, 1, 2, 3); method of diagnosis (histology, cytology, clinical, unknown). Tumour characteristics included: histological type (high-grade serous carcinoma, mucinous carcinoma, endometrioid carcinoma, clear cell carcinoma, carcinosarcoma, other); grade (G1, G2, G3); stage (II, III, IV). Missing data were allocated to a separate category. Treatment patterns were grouped into: chemotherapy then surgery; surgery then chemotherapy; surgery alone; chemotherapy alone; other/ unclear treatment; neither surgery nor chemotherapy.

Overall survival (OS) and progression-free survival (PFS) were examined by age and other factors, using Kaplan-Meier analysis for survival curves and Cox models for regression analysis, with time since diagnosis as the underlying time variable. Participants contributed follow-up time from the date of diagnosis until the earliest of: date of death, last date of follow-up, or date of recorded progression/ recurrence (for PFS only). Cox regression models were used to estimate hazard ratios (hereafter referred to as relative risks) for the association between age at diagnosis (75+ versus <75 years) and survival with adjustment for potential confounders. Stratification was used as an alternative to adjustment where there was concern regarding violation of the non-proportional hazards assumption (as assessed by tests based on Schoenfeld residuals). Quantitative methods were used to assess the magnitude of confounding, calculating the percentage change in the chi-squared statistic for the association between age and survival on adjustment/ stratification for one factor at a time. Analyses of PFS were censored at three years, as limited follow-up data were available beyond this date. Figures of Kaplan-Meier survival curves were censored at three years for both OS and PFS for simplicity of comparison, though regression analyses of OS used all available follow-up time. Death within three years was considered to be a progression event even if no prior progression was recorded (rather than a censoring event for PFS).

1. Townsend P, Phillimore P, Beattie A. Health and deprivation: inequality and the north. London: Croom Helm 1988.

2. Oken MM, Creech RH, Tormey DC, et al. Toxicity and response criteria of the Eastern Cooperative Oncology Group. *Am J Clin Oncol* 1982;5(6):649-55.

3. Piccirillo JF, Creech C, Zequeira R, et al. Inclusion of comorbidity into oncology data registries. *J Registry Manag* 1999;26(2):66-70.

**Table S1: Age distribution of patients with ovarian cancer by study site**

|  | **Age <75 years**  **(n=495)** | **Age 75+ years**  **(n=226)** | **All**  **(n=721)** |
| --- | --- | --- | --- |
| **Study site** |  |  | **p<0.001** |
| 1 | 46 (61%) | 30 (39%) | 76 (100%) |
| 2 | 67 (79%) | 18 (21%) | 85 (100%) |
| 3 | 123 (69%) | 54 (31%) | 177 (100%) |
| 4 | 120 (61%) | 77 (39%) | 197 (100%) |
| 5 | 47 (64%) | 27 (36%) | 74 (100%) |
| 6 | 92 (82%) | 20 (18%) | 112 (100%) |

Notes: Table shows row %; p-value is for Chi-2 test between study site and age.

**Table S2: Treatment of patients with ovarian cancer by age at diagnosis**

|  | **Age <75 years**  **N (%)** | **Age 75+ years**  **N (%)** | **All**  **N (%)** |
| --- | --- | --- | --- |
| **Treatment** |  |  | **p<0.0001** |
| **Surgery alone** | 21 (4%) | 9 (4%) | 30 (4%) |
| **Chemotherapy alone** | 78 (16%) | 55 (24%) | 133 (18%) |
| **Chemotherapy then surgery** | 175 (35%) | 39 (17%) | 214 (30%) |
| **Surgery then chemotherapy** | 145 (29%) | 33 (15%) | 178 (25%) |
| **Other/ unclear treatment** | 54 (11%) | 34 (15%) | 88 (12%) |
| **Neither surgery nor chemotherapy** | 22 (4%) | 56 (25%) | 78 (11%) |
| **Total** | 495 (100%) | 226 (100%) | 721 (100%) |

Notes: Table shows column %; p-value is for Chi-2 test between treatment and age.

**Table S3: Patient characteristics by age at diagnosis in those who received combination therapy (chemotherapy + surgery, in either order)**

| **Variable** | **Age <75 years**  **(n=320)** | **Age 75+ years**  **(n=72)** | **All**  **(n=392)** |
| --- | --- | --- | --- |
| **Site** |  |  | **p=0.09** |
| 1 | 31 (10%) | 13 (18%) | 44 (11%) |
| 2 | 54 (17%) | 5 (7%) | 59 (15%) |
| 3 | 73 (23%) | 19 (26%) | 92 (23%) |
| 4 | 47 (15%) | 10 (14%) | 57 (15%) |
| 5 | 43 (13%) | 13 (18%) | 56 (14%) |
| 6 | 72 (23%) | 12 (17%) | 84 (21%) |
| **Smoking** |  |  | **p=0.02** |
| Never (or not in last 10 years) | 192 (60%) | 51 (71%) | 243 (62%) |
| Past (in last 10 years) | 36 (11%) | 3 (4%) | 39 (10%) |
| Current | 32 (10%) | 1 (1%) | 33 (8%) |
| Unknown | 60 (19%) | 17 (24%) | 77 (20%) |
| **Index of Deprivation** |  |  | **p=0.5** |
| 1-5 | 180 (56%) | 38 (53%) | 218 (56%) |
| 6-10 | 135 (42%) | 34 (47%) | 169 (43%) |
| Unknown | 5 (2%) | 0 (0%) | 5 (1%) |
| **BMI** |  |  | **p=0.2** |
| <25 | 104 (33%) | 25 (35%) | 129 (33%) |
| 25-29 | 92 (29%) | 23 (32%) | 115 (29%) |
| 30+ | 89 (28%) | 12 (17%) | 101 (26%) |
| Unknown | 35 (11%) | 12 (17%) | 47 (12%) |
| **WHO Performance status** |  |  | **p=0.03** |
| 0 | 187 (58%) | 33 (46%) | 220 (56%) |
| 1 | 77 (24%) | 26 (36%) | 103 (26%) |
| 2+ | 31 (10%) | 11 (15%) | 42 (11%) |
| Unknown | 25 (8%) | 2 (3%) | 27 (7%) |
| **ACE-27** |  |  | **p=0.7** |
| 0 | 65 (20%) | 12 (17%) | 77 (20%) |
| 1 | 45 (14%) | 14 (19%) | 59 (15%) |
| 2 | 31 (10%) | 9 (13%) | 40 (10%) |
| 3 | 78 (24%) | 17 (24%) | 95 (24%) |
| Unknown | 101 (32%) | 20 (28%) | 121 (31%) |
| **Method of diagnosis** |  |  | **p=0.6** |
| Histology | 309 (97%) | 68 (94%) | 377 (96%) |
| Cytology | 9 (3%) | 3 (4%) | 12 (3%) |
| Clinical | 1 (0.3%) | 1 (1%) | 2 (0.5%) |
| Unknown | 1 (0.3%) | 0 (0%) | 1 (0.3%) |
| **Stage** |  |  | **p=0.9** |
| Stage 2 | 44 (14%) | 8 (11%) | 52 (13%) |
| Stage 3 | 202 (63%) | 47 (65%) | 249 (64%) |
| Stage 4 | 73 (23%) | 17 (24%) | 90 (23%) |
| Unknown | 1 (0.3%) | 0 (0%) | 1 (0.3%) |

Notes: p-values shown are for Chi-2 tests of association between age and each variable in turn.

**Table S4: Treatment for ovarian cancer by study site**

|  | **Site, N (%)** | | | | | | **p<0.0001** |
| --- | --- | --- | --- | --- | --- | --- | --- |
| **Treatment** | **1** | **2** | **3** | **4** | **5** | **6** | **Total** |
| **Surgery alone** | 1 (1) | 4 (5) | 6 (3) | 13 (7) | 1 (1) | 5 (4) | 30 (4) |
| **Chemo alone** | 18 (24) | 17 (20) | 0 (0) | 74 (38) | 10 (14) | 14 (13) | 133 (18) |
| **Chemo then surgery** | 20 (26) | 39 (46) | 52 (29) | 31 (16) | 33 (45) | 39 (35) | 214 (30) |
| **Surgery then chemo** | 24 (32) | 20 (24) | 40 (23) | 26 (13) | 23 (31) | 45 (40) | 178 (25) |
| **Other/ unclear** | 3 (4) | 5 (6) | 77 (44) | 3 (2) | 0 (0) | 2 (2) | 88 (12) |
| **Neither surgery nor chemo** | 10 (13) | 0 (0) | 2 (1) | 50 (25) | 7 (9) | 9 (8) | 78 (11) |
| **Total** | 76 (100) | 85 (100) | 177 (100) | 197 (100) | 74 (100) | 112 (100) | 721 (100) |

Notes: Table shows column %; p-value is for Chi-2 test for association between treatment and site.

**Table S5: Treatment for ovarian cancer by study site and age at diagnosis**

|  | **Site 1**  **(p=0.2)** | | **Site 2**  **(p<0.001)** | | **Site 3**  **(p=0.008)** | | **Site 4**  **(p<0.001)** | | **Site 5**  **(p=0.001)** | | **Site 6**  **(p=0.1)** | |
| --- | --- | --- | --- | --- | --- | --- | --- | --- | --- | --- | --- | --- |
| **Treatment** | **Age <75**  **N (%)** | **Age 75+**  **N (%)** | **Age <75**  **N (%)** | **Age 75+**  **N (%)** | **Age <75**  **N (%)** | **Age 75+**  **N (%)** | **Age <75**  **N (%)** | **Age 75+**  **N (%)** | **Age <75**  **N (%)** | **Age 75+**  **N (%)** | **Age <75**  **N (%)** | **Age 75+**  **N (%)** |
| **Surgery alone** | 1 (2) | 0 (0) | 3 (4) | 1 (6) | 5 (4) | 1 (2) | 9 (8) | 4 (5) | 0 (0) | 1 (4) | 3 (3) | 2 (10) |
| **Chemo alone** | 9 (20) | 9 (30) | 5 (7) | 12 (67) | 0 (0) | 0 (0) | 50 (42) | 24 (31) | 2 (4) | 8 (30) | 12 (13) | 2 (10) |
| **Chemo then surgery** | 14 (30) | 6 (20) | 35 (52) | 4 (22) | 42 (34) | 10 (19) | 27 (23) | 4 (5) | 24 (51) | 9 (33) | 33 (36) | 6 (30) |
| **Surgery then chemo** | 17 (37) | 7 (23) | 19 (28) | 1 (6) | 31 (25) | 9 (17) | 20 (17) | 6 (8) | 19 (40) | 4 (15) | 39 (42) | 6 (30) |
| **Other/Unclear** | 2 (4) | 1 (3) | 5 (7) | 0 (0) | 45 (37) | 32 (59) | 2 (2) | 1 (1) | 0 (0) | 0 (0) | 0 (0) | 0 (0) |
| **Neither surgery nor chemo** | 3 (7) | 7 (23) | 0 (0) | 0 (0) | 0 (0) | 2 (4) | 12 (10) | 38 (49) | 2 (4) | 5 (19) | 5 (5) | 4 (20) |
| **Total** | 46 (100) | 30 (100) | 67 (100) | 18 (100) | 123 (100) | 54 (100) | 120 (100) | 77 (100) | 47 (100) | 27 (100) | 92 (100) | 20 (100) |

Notes: Table shows column %.

P-values for each site are for Chi-2 tests for association between treatment and age within each site.

**Figure S1: Sankey diagram showing differences in treatment pathways for ovarian cancer by age at diagnosis**

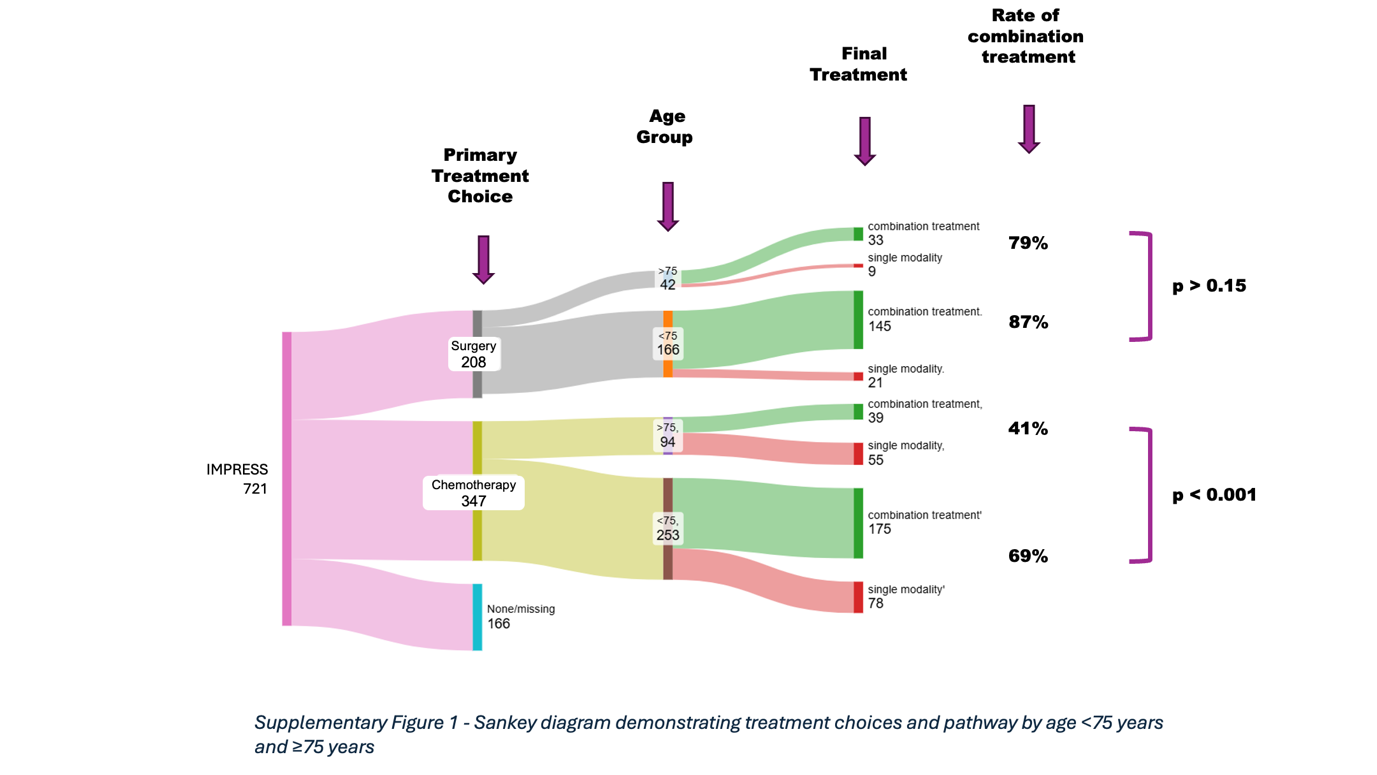

**Figure S2: Association between age at diagnosis (75+ versus <75 years) and overall survival (risk of death) with adjustment for various factors**
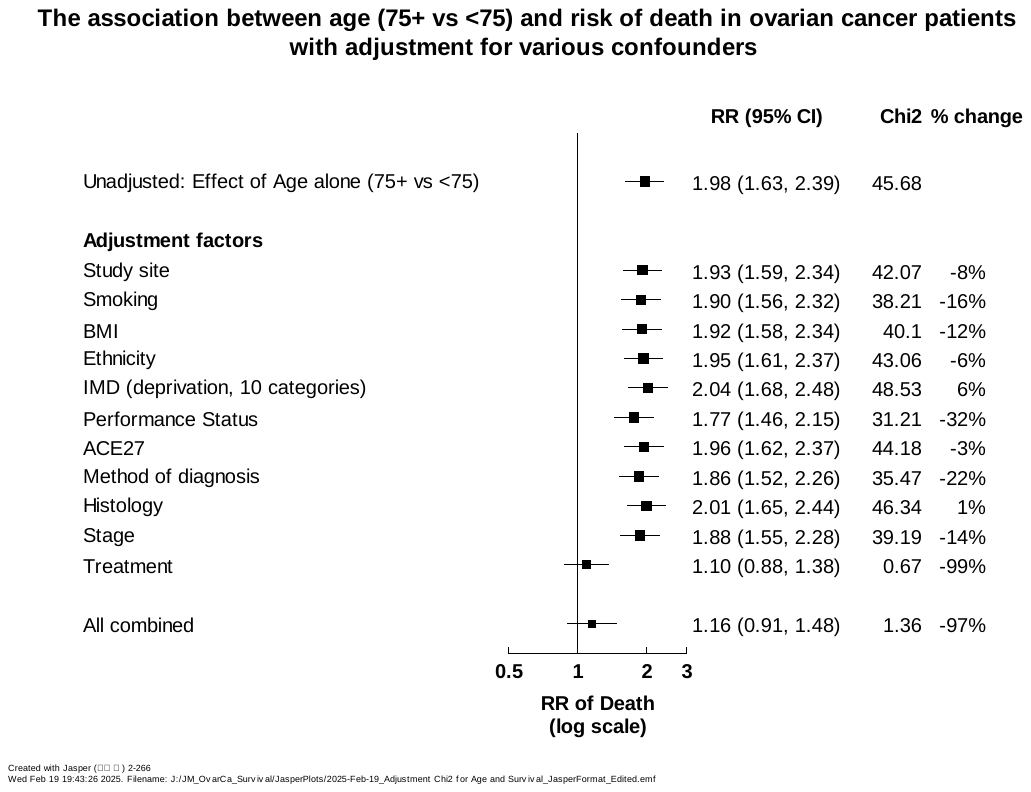

Notes: Figure S2 shows the relative risk of death (i.e. overall survival failure) from a Cox regression model, in women with ovarian cancer aged 75+ years at diagnosis versus <75 years, with adjustment for different potential confounders. Also shown are the Chi-2 statistic for the association, and the % change in Chi-2 statistic relative to the unadjusted model. Abbreviations: BMI, Body Mass Index; IMD, Index of Multiple Deprivation; ACE27, Adult Comorbidity Evaluation-27.

**Figure S3: Association between age at diagnosis (75+ versus <75 years) and overall survival (risk of death) with stratification for various factors**
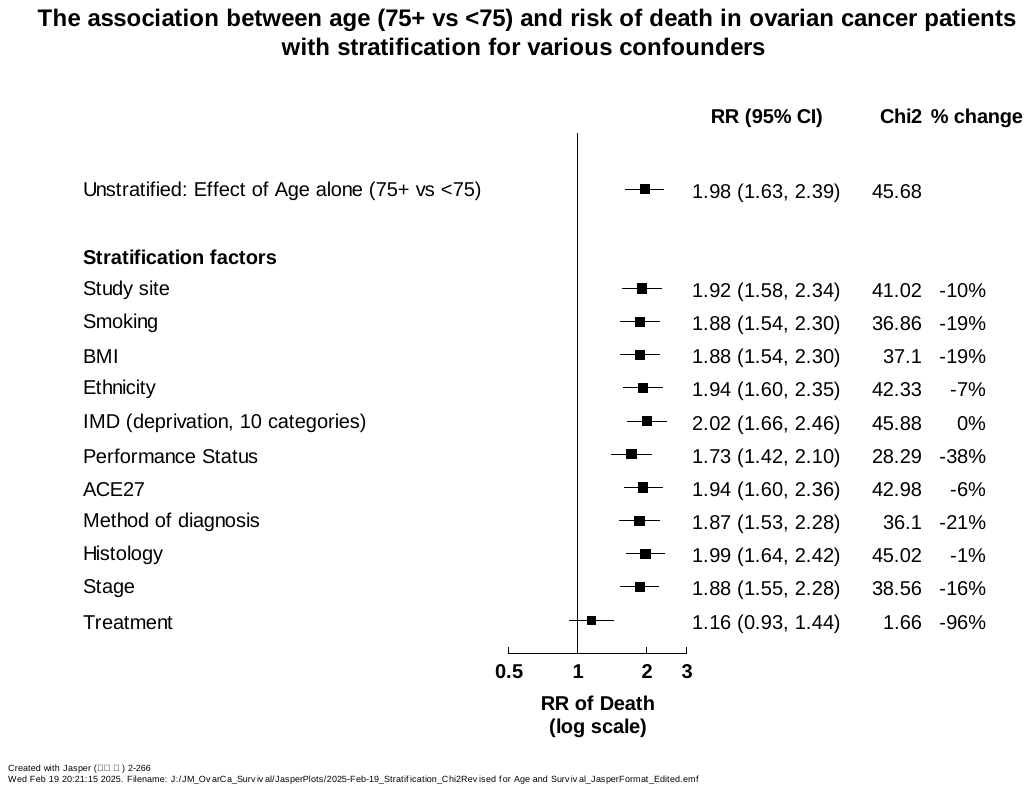

Notes: Figure S3 shows the relative risk of death (i.e. overall survival failure) from a Cox regression model, in women with ovarian cancer aged 75+ years at diagnosis versus <75 years, with stratification for different potential confounders. Also shown are the Chi-2 statistic for the association, and the % change in Chi-2 statistic relative to the unadjusted model. Abbreviations: BMI, Body Mass Index; IMD, Index of Multiple Deprivation; ACE27, Adult Comorbidity Evaluation-27.

**Figure S4: Association between age at diagnosis (75+ versus <75 years) and overall survival (risk of death) by treatment group**

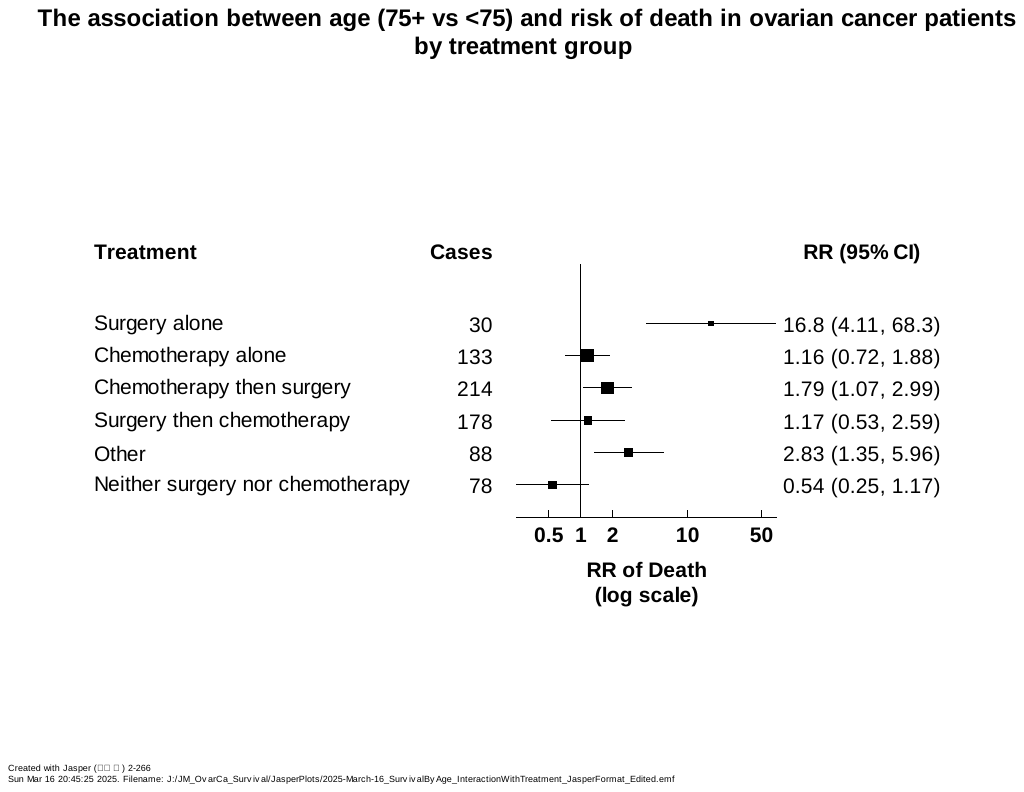

Notes: Figure S4 shows the relative risk (RR) of death comparing those aged 75+ at diagnosis vs aged <75 in different treatment groups. Results are from a Cox regression model for an interaction between age and treatment, stratified by study site, performance status, stage, and histological type, and adjusted for smoking, BMI, and method of diagnosis. Likelihood ratio test for interaction between age and treatment: p=0.0004.

**Figure S5: Kaplan-Meier graphs of overall survival by study site, performance status, stage at diagnosis, and histological type, shown separately for women aged <75 years versus 75+ years at diagnosis**

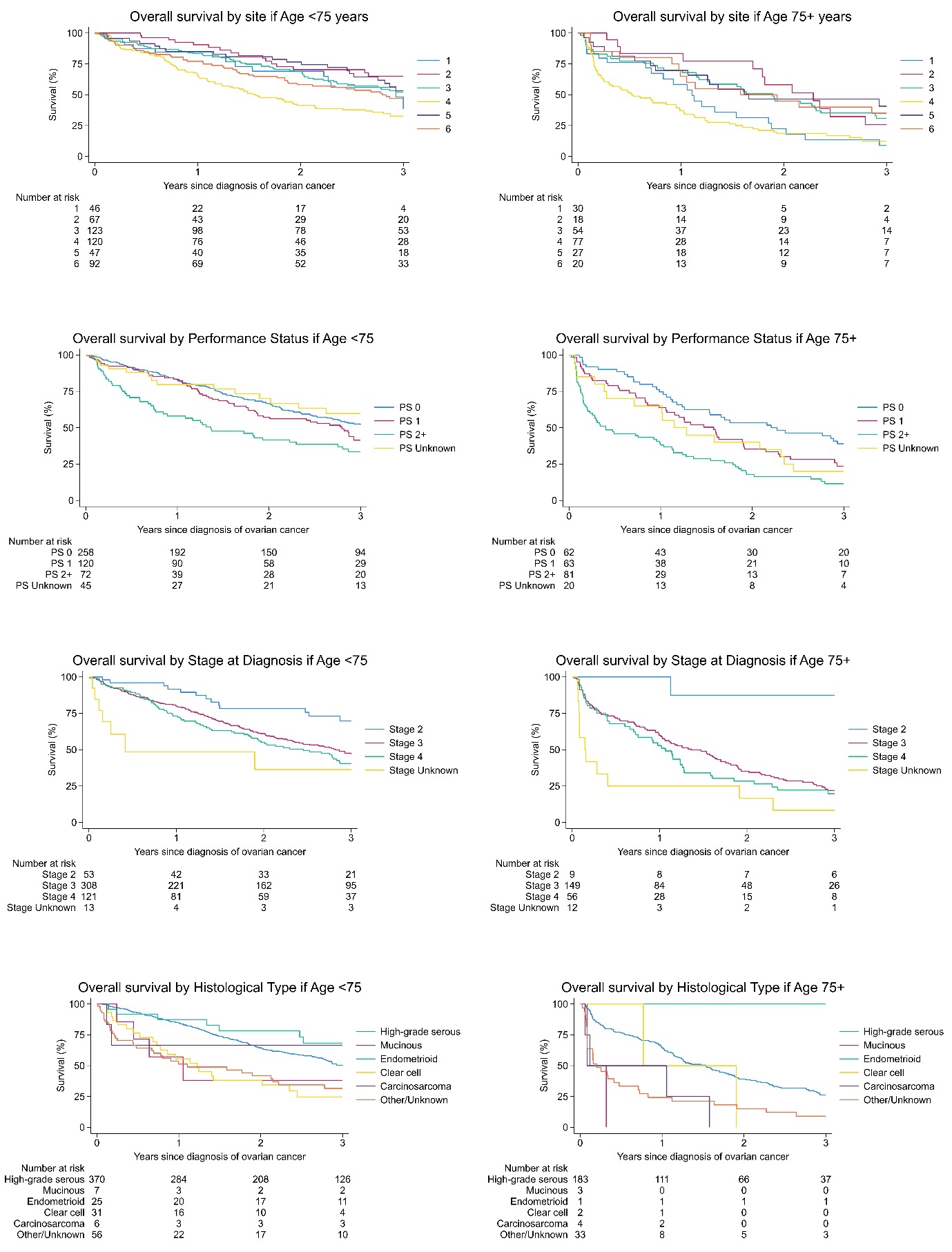

**Figure S6: Kaplan-Meier graphs of progression-free survival (PFS) by age at diagnosis and treatment**

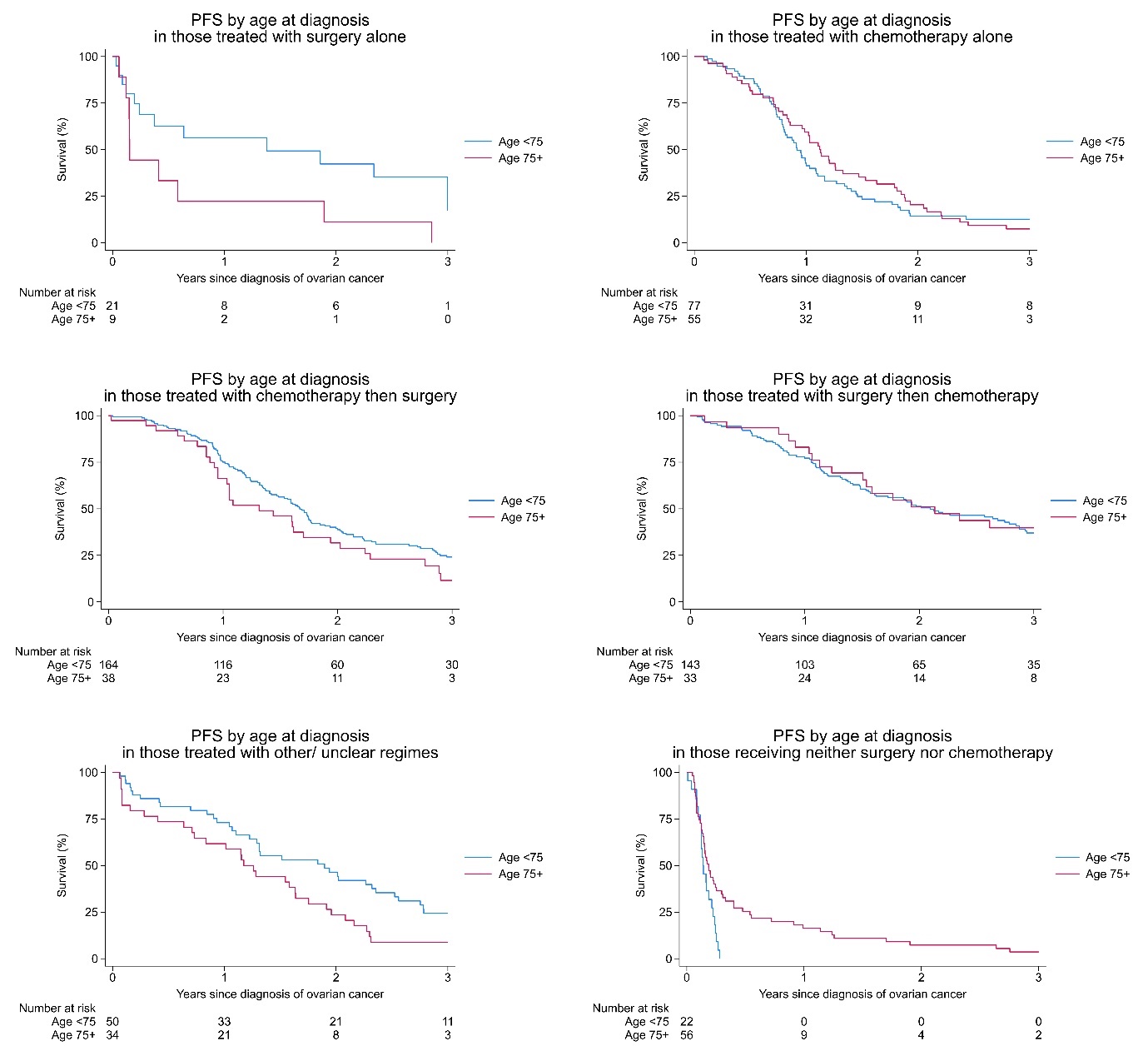

**Figure S7: Association between age at diagnosis (75+ versus <75 years) and progression-free survival (risk of progression or death) by treatment group**

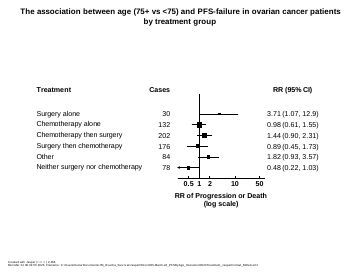
Notes: Figure S7 shows the relative risk (RR) of progression or death comparing those aged 75+ at diagnosis vs aged <75 in different treatment groups. Results are from a Cox regression model for an interaction between age and treatment, stratified by study site, performance status, stage, and histological type, and adjusted for smoking, BMI, and method of diagnosis. Likelihood ratio test for interaction between age and treatment: p=0.03.
